# Beyond intensity: Pain distribution shapes healthcare- and treatment-seeking beliefs in individuals with and without clinical pain

**DOI:** 10.64898/2026.04.02.26349577

**Authors:** Tom Frankenstein, Sophia Intert, Tibor M. Szikszay, Michał Katra, Bernhard Elsner, Robert C. Coghill, Kerstin Luedtke, Wacław M. Adamczyk

## Abstract

Pain is commonly described in sensory terms, yet its spatial characteristics—localization and distribution—are rarely quantified. We investigated whether lay beliefs about pain distribution influence theoretical decisions to seek care and treatment preferences. In a cross-sectional survey (N=503; 49% reporting pain), participants completed thought experiments in which both visually presented pain distribution patterns (small, moderate or large) and pain intensity (Numerical Rating Scale scores: 2, 5, 8/10) were systemically varied. For each scenario, they rated the likelihood of (i) seeking professional help (LoSH) and (ii) taking analgesic medication (LoTM). Participants also completed a spatial–intensity trade-off task, in which they chose between a fixed 20% reduction in intensity and variable reductions in pain distribution (20–80%). A reversed version contrasted a fixed 80% reduction in pain distribution with variable reductions in pain intensity. LoSH and LoTM increased significantly with greater pain distribution (*p*<0.001), mirroring the gradient observed for pain intensity. In the spatial–intensity trade-off task, participants’ choice followed a sigmoid-like function (*p*<0.001): 1% reduction in intensity was treated as equivalent to approximately a 3% reduction in distribution, indicating a systematic valuation of pain distribution. This ratio was lower in individuals experiencing pain compared to pain-free individuals. Moreover, 61% reported that pain distribution should be routinely considered in pain management alongside intensity. Results suggest that pain distribution is not merely a trivial descriptor, but a meaningful determinant of healthcare-related decision-making beliefs. Incorporating spatial metrics into clinical assessment and research may better capture how individuals implicitly evaluate pain severity.

**Summary:** Pain distribution (PD) influences healthcare and treatment decisions. Participants value PD reduction at approximately one-third of an equivalent decrease in pain intensity.

## 1. INTRODUCTION

Pain is inherently spatial, yet its spatial characteristics are rarely treated as a core dimension of pain assessment. However, beyond how much it hurts, pain is also defined by where it is and how it is distributed across the body [11,21,55]. Localization [72], as well as global [17,19] and local [3] patterns of pain distribution (PD), play a central role in diagnostic reasoning and may inform mechanistic inference [6,26,31,71]. Indeed, e.g. neuropathic pain often follows plausible neuroanatomical territories, whereas nociceptive pain tends to be more localized [71]. PD can change over time [66] like in complex regional pain syndrome, where pain can spread beyond the initial site to ipsilateral or contralateral body regions [49]. The concept of nociplastic pain further emphasizes widespread or diffuse PDs [38] as manifestation of altered central processing rather than peripheral tissue pathology [39]. Consistent with this, epidemiological evidence indicates that, compared with more local pain, widespread pain is associated with greater disability [30,78], reduced quality of life [34], and increased sick leave [18], underscoring the diagnostic and prognostic relevance of PD [69].

Neurophysiological evidence further supports the relevance of spatial characteristics of pain. Indeed, nociceptive processing is spatially distributed along the neuroaxis [13] and involves the convergence of afferent input [74], receptive field organization [13,35] as well as integration mechanisms such as spatial summation [2,14,59] and lateral inhibition [27,62,75]. Moreover, receptive field expansion and increased central excitability can enlarge PD [8,35], supporting the view that spatial pain characteristics have biologically grounded correlates. Collectively, these findings indicate that spatial features of pain are not merely descriptive but may provide a basis for clinical and mechanistic inference. Nevertheless, spatial information remains markedly underreported in pain research, where intensity continues to be prioritized over PD [43,67]. While spatial patterns guide diagnostic reasoning [70], it remains unclear how individuals interpret different distributions of pain when making healthcare-related decisions. PD may function as a severity-relevant cue, influencing perceived threat and driving behavioural responses.

Although PD has been associated with increased healthcare utilization [54], isolated investigation of how PD patterns shape individuals’ decisions to seek care and initiate treatment is lacking. Clarifying this relationship may inform the development of more personalized approaches to pain management. Thus, to address this problem, the present study employed structured thought experiments that independently manipulated PD and pain intensity. We examined whether spatial extent influences hypothetical judgments regarding likelihood of seeking professional care and taking analgesic medications. In addition, a new task was developed and used to determine whether individuals exhibit a quantifiable exchange rate between reductions in pain intensity and reductions in spatial extent. We hypothesized that (i) greater PD would increase the likelihood of seeking help and medication use and (ii) that this effect would be more pronounced in individuals experiencing pain. We further predicted that (iii) higher pain intensity would produce similar effects as larger PD. Finally, we aimed to explore if individuals would demonstrate a systematic trade-off between intensity and PD, indicating that PD is weighted as a severity-relevant dimension of pain.

## 2. MATERIAL AND METHODS

### 2.1. General information

In this cross-sectional nationwide online study conducted in Germany, participants completed a series of thought experiments and additional questions assessing their beliefs regarding spatial aspects of pain. Data collection began in February 2025 and continued until the a priori target sample size of 500 responses was reached in February 2026 (**Appendix S1**). The study protocol was approved by the local Ethics Committee of the University of Lübeck (No. 2025-044 from 28.01.2025 / CL). The study design and analysis plan were preregistered on the Open Science Framework using the AsPredicted.org template (https://osf.io/cvjdm/overview). Following survey development, the instrument was piloted in a sample of 8 participants. Based on pilot feedback, minor amendments were made to the study structure, including adjustments to plausibility checks **(Appendix S2)**. Participation was fully voluntary, and no financial incentives were provided thus minimising the potential risk of survey being a target for AI-bots. The study is reported in accordance with the Checklist for Reporting Results of Internet E-Surveys (CHERRIES) [24]. A transparency statement detailing all deviations from the preregistered study protocol can be found in the **Appendix** (**S3**).

### 2.2. Recruitment and sampling

Adults aged 18 years or older were eligible to participate, irrespective of current pain status. Recruitment was conducted via announcements on social media platforms (e.g., Facebook, Instagram), professional networks, and online forums using a convenience sampling approach. The study flyers were additionally distributed in printed form on the campus of the University of Lübeck, at the University Medical Centre and medical institutions acting as partners of the University. To recruit individuals with pain, we contacted pain networks and patient organizations (e.g., patient organizations of the German Pain Society). No crowdsourcing recruitment platforms (e.g., Prolific or Amazon Mechanical Turk) were used. Participation involved minimal risk and no anticipated adverse effects. No personally identifiable information was collected. All data were stored and analyzed in anonymized form, with each participant assigned a unique study identification number.

### 2.3. Sample size

A priori sample size considerations were based on standard parameters for descriptive studies. Assuming a margin of error of 5%, maximum variability (p = 0.5), and a 95% confidence level (Z = 1.96), an estimated minimum of 385 participants was required for adequate descriptive precision (http://www.openepi.com/SampleSize/). To account for potential missing data and to enable planned subgroup analyses (e.g., by pain status), the recruitment target was set at a minimum of 500 participants. Such sample size was also expected to provide sufficient statistical power to detect potentially small effects of PD in the primary analyses. No upper limit on participant numbers was predefined.

### 2.4. Study design

This study employed a cross-sectional design, with data collected at a single time point via an online questionnaire. The questionnaire was developed specifically for this study (in German) and included closed-ended items (multiple-choice and scale-based responses) as well as open-ended questions. For sociodemographic variables requiring numerical input (e.g., height, weight), response formats were predefined within the survey settings prior to data collection (e.g., MM.YYYY for dates), thereby promoting data consistency and reducing entry errors. The survey was hosted on the SoSci Survey platform (https://www.soscisurvey.de/), a web-based research tool developed in Germany (SoSci Survey GmbH, Munich, Germany). The platform provided secure data handling and complied with applicable data protection regulations.

### 2.5. Survey structure

Upon accessing the survey, participants were presented with an introductory information page outlining the purpose and scope of the study. The text explained that pain comprises multiple dimensions and highlighted the distinction between the well-established intensity dimension (e.g., commonly measured using 0–10 Numerical Rating Scales, NRSs) and the comparatively understudied spatial dimension of PD. The study aim - to explore individuals’ perspectives on the spatial aspects of pain and how PD influences decision-making - was explicitly stated. It was clarified that the questionnaire was open to all adults, regardless of whether they were currently experiencing pain or not. It should be noted that while the introduction provided participants with a detailed overview of the survey’s scope and thematic focus, it was intentionally written avoiding technical terminology - such as “spatial-intensity trade-off task” - within the participant-facing instructions. This was a deliberate design choice aimed at minimizing potential framing effects, ensuring that participants’ responses were based on an intuitive evaluation of the pain scenarios rather than an understanding of the underlying experimental mechanism. Participants were informed that completion would take approximately 10–15 minutes, that the survey included both multiple-choice and open-ended questions, and that additional items would be presented to those reporting current pain. The initial page also provided information that participation was described as involving no risks or negative consequences. It was stated that no identifying personal data would be collected and that all results would be analyzed and published in anonymized form. Finally, detailed data protection information was provided in accordance with the General Data Protection Regulation. This included the legal clause for data processing (personal consent), procedures for pseudonymization, data storage location, restrictions on data transfer, rights of withdrawal, and contact details of both the responsible investigator and the institutional data protection officer. Proceeding to the questionnaire by clicking “Next” constituted informed consent to participate. The survey itself composed of five distinct sections, which are described below.

#### 2.5.1. Sociodemographic section

In this initial section, demographic and anthropometric data were collected to characterize the study sample. Participants reported their year of birth, from which age was derived, as well as sex assigned at birth and laterality. Height (cm) and weight (kg) were recorded as continuous variables. Highest educational attainment was assessed using a predefined categorical list of response options. Physical activity was assessed by asking participants to provide a numerical estimate of the average time spent engaging in any form of physical exercise per week (in minutes). Because the survey involved the visual presentation of PD patterns, participants were also asked to indicate the type of device used to complete the survey, categorized by screen size (smartphone, tablet, or computer screen), in order to control for potential effects related to display characteristics.

#### 2.5.2. Thought experiments with pain distribution

In this (main) section, participants were presented with a series of structured thought experiments based on hypothetical distributions of pain. Three distinct unilateral PD patterns were displayed: small (e.g., limited to the foot), moderate (covering approximately half of the lower limb), and large (involving the entire lower limb). Participants were instructed to imagine that they had been experiencing pain in the displayed and highlighted area on a body-chart **(Figure 1)** for approximately 3–4 weeks. For each distribution pattern, they responded to two questions using an electronic Visual Analogue Scale (eVAS) with the left anchor set as “very unlikely” (0) and right one as “very likely” (100): (i) the likelihood of seeking help (LoSH) from a healthcare professional and (ii) the likelihood of taking analgesic medications (LoTM) given the depicted pain distribution **(Appendix S4** and **S5)**.

**Figure 1.**
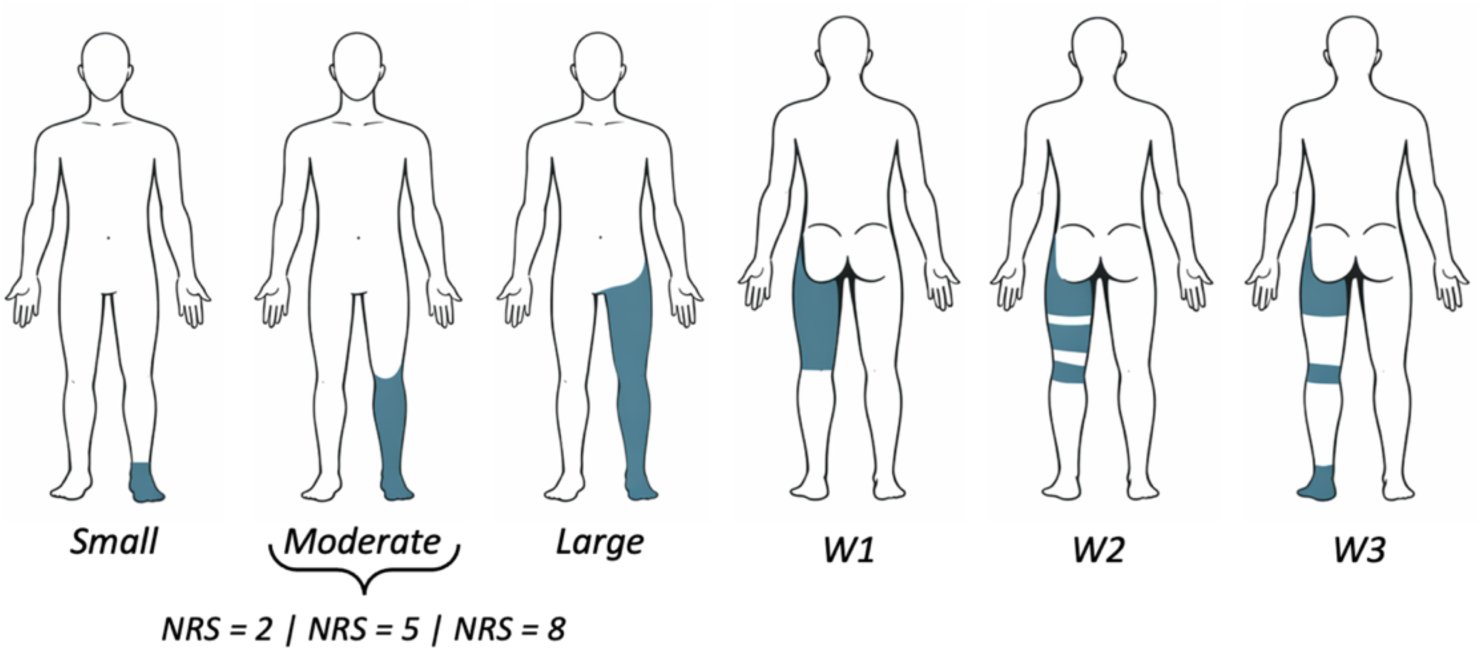
Design of thought experiments. Participants were presented with three levels of pain distribution (small, moderate, and large) across three body regions: lower limb, lower back, and upper limb. The moderate pain distribution was additionally presented at different pain intensity levels to allow direct comparison of the relative impact of pain distribution versus pain intensity on the primary outcomes: self-reported likelihood of seeking help (LoSH) and taking analgesic medication (LoTM). Body charts with widespread (W) patterns were labelled as W_1_ (contiguous, control), W_2_ (discrete, shrunk), and W_3_ (discrete, expanded) and were presented only for the lower limb. Pattern W_3_ was also presented across three intensity levels. Note: For illustrative purposes, this figure depicts only the lower limb condition as an example of the experimental stimulus set.

The three distribution sizes (small, moderate, large) were presented across three anatomical regions: upper limb, lower back, and lower limb. To enable comparison with pain intensity the same two questions were repeated for a moderate distribution pattern **(Figure 1)** while varying intensity at three levels: low (equivalent to 2/10 on NRS), moderate (5/10 NRS), and high (8/10 NRS). Finally, participants were presented with variations of widespread pain patterns limited to the lower limb only. To reduce the total duration of participation in the survey, only the LoSH was assessed in this condition, as it was considered the primary outcome of interest in this condition. In this scenario, pain previously presented as a moderate lower limb distribution was modified to represent three spatial configurations: contiguous, discrete-shrunk, and discrete-expanded **(Figure 1)**. In total, participants evaluated two response types (LoSH and LoTM) across three spatial extents and three intensity levels in three anatomical regions, with additional items examining widespread pain patterns (LoSH only).

#### 2.5.3. Beliefs about spatial aspects of pain

In the third section, participants completed five mandatory items assessing beliefs about the clinical significance of spatial aspects of pain. The first four items were rated on a five-point Likert scale ranging from −2 (strongly disagree) to +2 (strongly agree). These items assessed: (i) whether a larger spatial extent of pain reflects a more severe underlying injury, (ii) whether pain perceived as deeper within the body indicates a more serious or dangerous condition, (iii) whether pain management should target not only pain intensity but also the PD, and (iv) whether longer-lasting injuries are typically associated with a broader or more widespread PD. The fifth item was assessed using a bipolar semantic differential scale ranging from −5 to +5. Participants indicated whether they believed that greater spatial extent of pain (left anchor) or higher pain intensity (right anchor) is more indicative of a serious underlying injury.

#### 2.5.4. Spatial-intensity trade-off task (SITT)

Following the belief items, participants completed a SITT designed to estimate the relative subjective value assigned to reductions in pain intensity versus reductions in PD. The SITT was conceptually modelled on forced-choice paradigms used to estimate indifference points between competing attributes in behavioural economics and psychophysics [33,65]. Participants were instructed to imagine experiencing ongoing pain and were presented with a series of dichotomous forced-choice scenarios. In the first set of items, they chose between (i) a 20% reduction in pain intensity with no change in spatial extent and (ii) no reduction in pain intensity but a 20% reduction in the spatial extent of pain. Across successive items, the magnitude of the potential reduction in spatial extent increased in 10% increments, ranging from 20% to 80%, while the reduction in pain intensity remained constant at 20%. In the second set of items, the reference condition was reversed. Participants chose between (i) an 80% reduction in spatial extent with no change in intensity and (ii) no reduction in spatial extent but an 80% reduction in pain intensity. In these items, the magnitude of the potential reduction in pain intensity decreased in 10% increments from 80% to 20%, while the spatial reduction remained fixed at 80%. All items employed binary response options. Conceptually, the indifference point was defined as the point at which an individual values reduction in pain intensity and pain distribution as equal, such that neither option is clearly preferred over the other. The incremental manipulation of one dimension while holding the other constant enabled identification of the point (value in one domain) at which participants shifted their preference from one pain attribute to the other.

#### 2.5.5. Characterization of participants with pain

The final section of the survey was presented only to participants who indicated that they were <u>not</u> pain-free using a screening item (“are you currently pain-free?” yes/no). Participants who reported being pain-free exited the survey at this stage. Those reporting pain completed an additional module consisting of items characterizing current and recent pain status. Participants indicated whether they were experiencing pain at the moment of participation and whether they had experienced pain within the past 30 days (response options: yes, no, unsure). Current pain intensity and average pain intensity over the previous week were assessed using a NRS ranging from “no pain” (0) to “worst pain imaginable” (100). Pain localization was assessed using a multiple-choice item with predefined categories (upper limb, lower limb, upper spine, lower spine, head, other). If “other” was selected, participants were able to specify the location in free-text (open) format. To assess pain duration, participants had to recall and report the onset date of their pain. Chronicity was further characterized by asking whether pain had been present for approximately 50% of the time, more than 50%, or less than 50% during the previous six months [16,76,77]. Participants were asked whether they had received a medical diagnosis related to their pain (yes/no). If yes, they selected from 17 predefined diagnostic categories based on the International Classification of Diseases, 10th Revision (ICD-10), with an additional option allowing specification of unlisted diagnoses via free-text response – results from this question are not part of this report. Finally, participants indicated the spatial extent of their own pain using a multiple-choice item comprising five predefined categories reflecting increasing distribution size: pain extent of the size of finger (1), fist (2), hand (3), two hands (4), larger than two hands (5).

### 2.6. Data processing and statistical analysis

Raw survey data from participants who reached the last page of the survey were first cleaned and recoded (if necessary) for readability prior to analysis. For instance, invalid entries for date of birth or onset of pain were removed, whereas the correct entries and timestamps were used to calculate age and duration of pain, respectively. Although the survey did not offer any financial profit and participation was purely voluntary, thus minimising the risk of interference from autonomous internet programs “bots” [45], the survey results were inspected for potential bot activity and response patterns using three complementary indicators of completion time. First, the total completion time (TCT) in minutes was calculated for each respondent and distribution was visualized using probability density function and histograms. TCT reflects each participant’s absolute time spent on the survey, corrected for interruptions (page dwell times exceeding the expected range due to a paused session are replaced with the median dwell time of other respondents for that page). A conservative threshold of 1/3 of the median completion time was applied to flag unusually rapid respondents.

Second, a relative speed index (RSI) was computed for each participant as the ratio of the median completion time of the total sample to the completion time of the respective participant, with values above 1 indicating a faster-than-typical pace and values below 1 a slower pace [44]. Unlike TCT, which is an absolute duration, RSI expresses each participant’s pace relative to the cohort, which makes it comparatively insensitive to differences in absolute survey length between subgroups (e.g., participants completing additional condition-specific items). A recommended cut-off value of 2 was applied to flag potentially low-quality responses [44]. The resulting indices were visualized and compared between participants with and without pain (**Appendix S6**). Finally, the time spent on each page of the main survey section (pages 4–9) was analyzed separately, and the distribution of response times per page was visualized using density plots, allowing inspection of pacing irregularities at a finer grain than the whole-survey indicators above. These analyses collectively enabled the identification of potential anomalous response patterns indicative of non-human or inattentive participation.

Depending on the level of measurement and distribution of the variables, descriptive statistics were used to characterize the studied sample: For approximately normally distributed metric variables, means and standard deviations (SDs) were calculated. For ordinal or non-normally distributed variables, medians and interquartile ranges were used. Categorical variables were summarized using absolute frequencies and percentages. All data handling and analyses were conducted in R (version 4.6.1 [2026.06.24]) via RStudio (version 2026.06.0+242) [63]. Primary analyses addressing <u>hypothesis 1</u> in LoSH and LoTM were performed by using Linear Mixed Models (LMM) via “lme4” package. The “subject” was used in each model as a random factor whereas the primary factor “size” of the PD (small, moderate, large) was set as fixed. Additional fixed factor “site” (back, lower limb, upper limb) was set to explore potential interaction: LoSH/LoTM ∼ “site” × “size” + (1|“subject”). Fixed effects were reported using Satterthwaite approximation of degrees of freedom and *F* tests [41], and effect size measure (partial eta squared, *η^2^_p_*) was approximated via “effectsize” package according to the formula: “*F* × df_effect_) / (*F* × df_effect_ + df_error_)”. These analyses were repeated with “intensity” (2, 5, and 8/10) set as a fixed factor, using the same model used in the PD domain. The subject was modelled only as a random intercept, because models with random slopes for “size” or “intensity” (e.g., 1 + “size” | “subject” or 1 + “size” || “subject”) did not converge, resulting in a singularity issue. As random-intercept-only models may carry an elevated Type I error risk for within-subject fixed effects [7], all reported contrasts were subjected to *p*-value correction via False Discovery Rate (FDR).

Three patterns of widespread pain **(Figure 1)** were analysed separately with distinct LMM as the level of this factor did not have a graded structure as in other conditions. All significant main and interaction effects were followed by post-hoc comparisons. To address <u>hypothesis 2</u> and thus compare the influence of PD and pain intensity on health-care seeking judgements, we modelled LoSH and LoTM as a function of predictor type (size vs. intensity), pain level (low, interim, high), and their interaction, using LMM with random intercepts per participant: LoSH/LoTM ∼ “predictor” × “level” + (1 | “subject”). To evaluate the appropriateness of the linear mixed-effects models for the bounded 0–100 outcome measures, we assessed clustering at the scale boundaries (0 and 100) (**Appendix S7**). Additional analyses encompassed testing for the main effects of “pain status” (fixed factor) defined by the results of the questions about the current pain (yes, no), and interaction of this factor with “size”/“intensity”. This analysis followed the same logic as described above and was primary used to address <u>hypothesis 3</u>.

The spatial-intensity trade-off task data were analyzed using Generalized Linear Mixed Models (GLMM) via “lme4” package with binary response variable being “selected PD” (0, 1 = yes) using the formula with distribution set to binomial: response ∼ “trade level” (20:20% to 20:80%) + (1|“subject”). The model was repeated by adding between-subject factor “pain status” (having vs. not having pain) and also with and without confounders (age, weight, education, device used). Subject-level ratios of PD selection over pain intensity were extracted and also compared between individual with and without pain using *t* tests (Welch). Between-group differences in descriptive variables were calculated using unpaired *t* tests, χ^2^, Mann-Whitney *U*, or Fischer’s exact tests.

In addition, post hoc power analyses were conducted to contextualize the observed effect sizes (**Appendix S8**). Responses from open question (site of pain) were analyzed using thematic analysis to identify recurring themes/codes and patterns [10]. Responses were independently reviewed by two authors (TF and WA) and, where possible, assigned to the existing body-area categories. Discrepancies were resolved by a third reviewer (TS). During this process, an additional category, “whole-body pain,” was created to accommodate responses that could not be meaningfully assigned to the predefined categories. Due to the small number of participants in the “whole-body pain” category, no subgroup analyses were performed.

## 3. RESULTS

A total of 688 individuals initiated the survey, and 503 completed it (49% reported pain), yielding a completion rate of 73.1%. The average time to complete the survey was 13 min (SD ± 6). Data completeness in the final analytic dataset was high: across variables, the proportion of missing values ranged from 0% to 8.47%. When quantified at the cell level, the dataset contained 8523 eligible data cells, of which 82 were missing, corresponding to an overall completeness of 99.04% with the used dataset.

The distribution of completion times among participants showed expected patterns **(Figure 2)**. Only a small number of respondents (*n* = 4) completed the survey in less than 1/3 of the median completion time, and a minority exhibited RSI exceeding 2. Density plots of response times across the main survey section (pages 4 to 9) and TCT revealed rather right skewed distribution with no clear deviations suggestive of automated participation neither within the entire cohort nor within the subgroups (pain vs. pain-free). Due to the small number of participants with high speed rates of completing the survey, and lack of a priori criteria for removal, main analyses were performed with retaining the whole study sample. Descriptive statistics with distinction into subgroups (pain vs. pain-free) is presented in **Table 1**. Clinical characteristics are also presented in **Table 2**.

**Figure 2.**
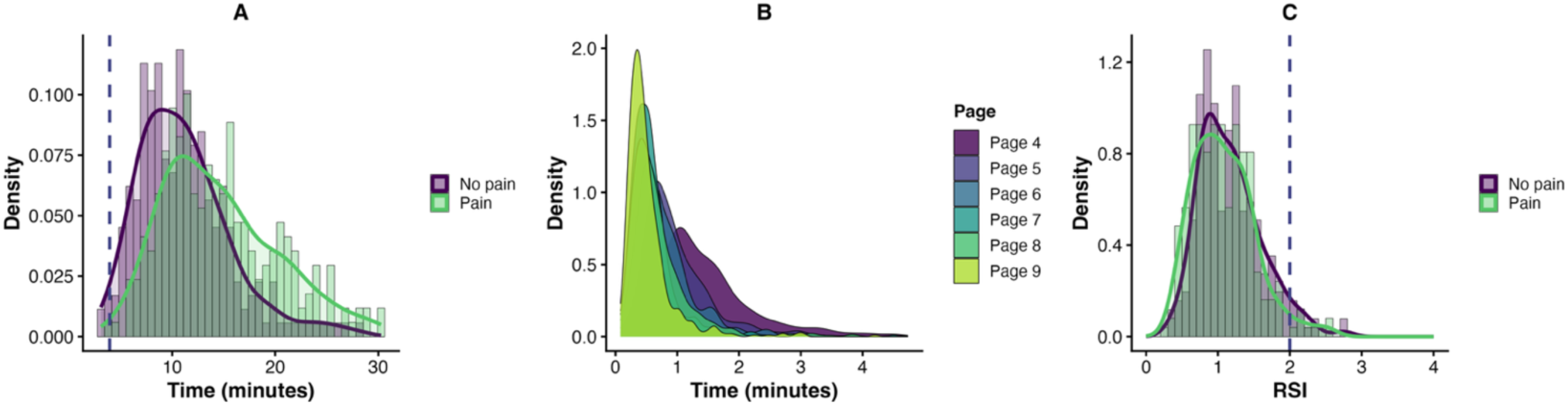
Veracity and quality of the survey’s data. <u>Left:</u> Histogram and density curve of the total completion time. The dashed vertical line points to 1/3 of the median completion time. Highlighted bar to the left from the line refers to four participants with completion score below 3:56s. <u>Middle:</u> Distribution of the Relative Speed Index (RSI) across participants - RSI is a ratio of sample’s median to individuals’ completion’s time. Dashed vertical line shows the cut-off threshold of 2 (two times faster than median). <u>Right:</u> Overlaid density distributions of the time spent on main survey sections which were similar but referred to different body regions (pages from 4 to 9). The curves visualize speed consistency across these pages indicating typical for humans learning pattern.

**Table 1.** Descriptive statistics.

| Variable | Estimate | Units | Pain-free | | Pain | | Statistic | $p$ value |
| --- | --- | --- | --- | --- | --- | --- | --- | --- |
|  |  |  | N |  | N |  |  |  |
| Age | $\bar{x}$, $SD$ | years | 255 | 34.13 (14.81) | 247* | 41.72 (16.43) | $t = -5.43$ | < 0.001 |
| Height | | cm | 252 | 172.15 (12.37) | 239 | 171.97 (8.42) | $t = 0.18$ | 0.85 |
| Weight | | kg | 251 | 70.92 (13.36) | 240 | 76.72 (17.13) | $t = -4.18$ | < 0.001 |
| PA | | min/week | 252 | 186.56 (164.18) | 232 | 169.54 (198.99) | $t = 1.02$ | 0.31 |
| Sex | $N$, % | male | 90 | (35%) | 69 | (28%) | $\chi^2 = 2.91$ | 0.09 |
|  |  | female | 165 | (65%) | 179 | (72%) |  |  |
| Laterality | | left | 20 | 8% | 17 | 7% | $\chi^2 = 0.06$ | 0.81 |
|  |  | right | 235 | 92% | 230 | 93% |  |  |
| Device used | $N$, % | phone | 141 | 56% | 163 | 66% | $\chi^2 = 6.20$ | < 0.05 |
|  |  | tablet | 29 | 12% | 29 | 12% |  |  |
|  |  | computer | 80 | 32% | 55 | 22% |  |  |
| Education | $N$, % | no formal | 0 | 0% | 2 | 1% | | < 0.001 |
|  |  | lower secondary | 7 | 3% | 47 | 19% |  |  |
|  |  | upper secondary | 111 | 44% | 85 | 34% |  |  |
|  |  | tertiary | 127 | 50% | 105 | 43% |  |  |
|  |  | doctoral degree | 8 | 3% | 8 | 3% |  |  |

Legend: SD – standard deviation, PA – physical activity. \*Participants with pain constituted 49% of the sample.

**Table 2.** Pain-related characteristics of participants reporting pain.

| Variable | Estimate | Units | N | Estimate |
| --- | --- | --- | --- | --- |
| Pain intensity (C) |  | NRS | 247 | 30.00 (24.63) |
| Pain intensity (W) | $\bar{x}$, $SD$ | NRS | 248 | 38.79 (22.46) |
| Pain duration |  | Months | 227 | 57.61 (102.59) |
| Pain last month | $N$ (%) | Yes | 248 | 145 (58.47) |
|  |  | No |  | 48 (19.35) |
|  |  | Unsure |  | 55 (22.18) |
| Pain Frequency | $N$ (%) | > 50% | 247 | 123 (49.80) |
|  |  | ~ 50% |  | 31 (12.55) |
|  |  | < 50% |  | 93 (37.65) |
| Pain during survey | $N$ (%) | Yes | 248 | 178 (71.77) |
|  |  | No |  | 70 (28.23) |
| Pain locations | $N$ (%) | Lower back | 248 | 102 (25.31) |
|  |  | Lower limb | 248 | 94 (23.33) |
|  |  | Upper limb | 248 | 71 (17.62) |
|  |  | Upper back | 248 | 55 (13.65) |
|  |  | Head | 248 | 54 (13.40) |
|  |  | Whole body pain | 248 | 7 (1.74) |
|  |  | Other | 248 | 20 (4.96) |
| Pain extent (size) | $N$ (%) | Finger | 248 | 20 (8.06) |
|  |  | Fist | 248 | 78 (31.45) |
|  |  | Hand | 248 | 66 (26.61) |
|  |  | = Two hands | 248 | 45 (18.15) |
|  |  | > Two hands | 248 | 39 (15.73) |
| | $M$, $IQR$ | Score | 248 | 3 (2 to 4) |

Legend: NRS – Numeric Rating Scale, SD – standard deviation, IQR – interquartile range, C – current pain, W – overall pain from last week, “Pain frequency”: pain experienced on more than, less than, or approximately 50% of days in a typical month. “Pain locations”: body region affected by pain (multiple choice question), “Pain extent (size)”: spatial size of the painful area, selected from five predefined options: pain of the size of: finger, fist, hand, two hands, or area > two hands.

### 3.1. Questions regarding the spatial aspects of pain

No differences were observed between individuals with and without pain in their responses to the five questions addressing the meaning and importance of spatial aspects of pain, thus responses were presented for entire sample **(Table 3)**.

**Table 3.** Mann–Whitney U comparisons between groups for Questions 1–5.

| Question | Pain-free, Md [IQR] | Pain group, Md [IQR] | U | pFDR | rrb [95% CI] |
| --- | --- | --- | --- | --- | --- |
| Q1 | 1 [-1, 1] | 0 [-1, 1] | 33 375 | 0.37 | 0.06 [-0.04, 0.16] |
| Q2 | 1 [0, 1] | 1 [0, 1] | 33 772 | 0.26 | 0.07 [-0.03, 0.17] |
| Q3 | 1 [0, 1] | 1 [0, 1] | 30 637 | 0.69 | -0.03 [-0.13, 0.07] |
| Q4 | 1 [-1, 1] | 1 [0, 1] | 29 177 | 0.26 | -0.07 [-0.17, 0.03] |
| Q5 | -3 [-4, -3] | -4 [-4, -3] | 33 248 | 0.38 | 0.06 [-0.05, 0.16] |

Note: Md = median, IQR = interquartile range, U = Mann–Whitney U statistic, pFDR = False Discovery Rate–corrected p value, rrb = rank-biserial correlation. Group sizes for all comparisons were n = 248 (pain group) and n = 254 (no-pain group). None of the group comparisons reached statistical significance after FDR correction.

Across the entire sample, 49% of participants agreed -to some extent- that a larger pain extent indicates a more severe injury (Q1), 70% agreed that deeper pain signals greater severity (Q2), 61% indicated that pain treatment should consider reduction of pain area in addition to pain intensity (Q3), and 57% agreed that more progressive injuries are associated with larger painful areas (Q4). When directly asked which dimension better signals a more serious injury (Q5) (larger area vs. higher intensity), 91% of respondents selected higher intensity **(Figure 3)**.

**Figure 3.**
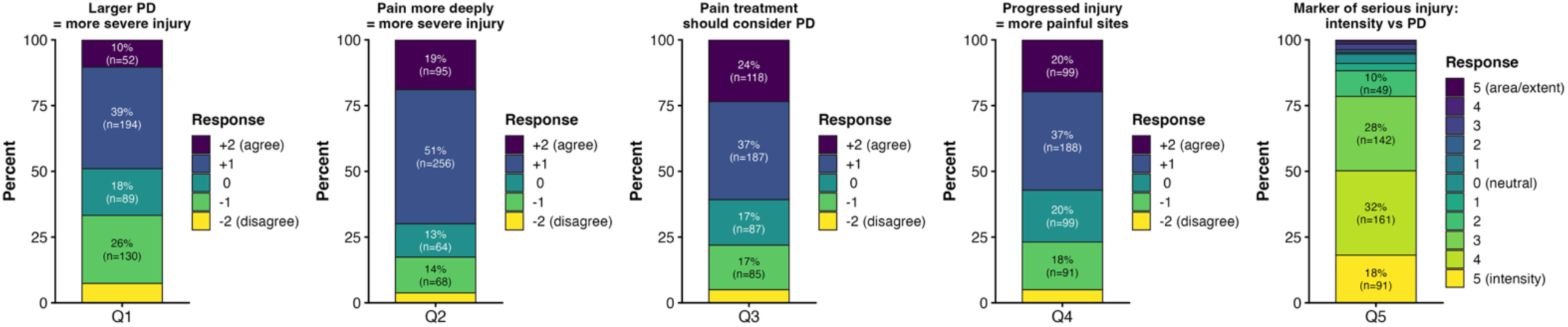
Responses to questions regarding the meaning of spatial aspects of pain. 61% of participants agreed or rather agreed that pain management should address pain distribution in addition to pain intensity. Mann–Whitney U tests revealed no significant differences between two subgroups of individuals: with and without pain in any of the questions (p ≥ 0.26).

### 3.2. Thought experiments within LoSH and LoTM

The primary LMM on LoSH revealed a significant “PD” × “site” interaction (*F*_(4, 4016)_ = 8.53, *p* < 0.001, *η_p_*² = 0.01), indicating that the effect of PD ratings varied across body regions. When PD was small, LoSH ratings for the lower limb were comparable to those for the lower back (*p* = 0.28). However, as PD increased, LoSH ratings formed the gradient: ratings were significantly higher for upper limb compared to the lower limb (*p* < 0.001), and for lower limb compared to the back (*p* < 0.001). Main effect of “PD” was also significant (*F*_(2, 4016)_ = 681.77, *p* < 0.001, *η_p_*² = 0.25, **Figure 4**), indicating that LoSH increased as the PD increased: small vs. moderate (*p* < 0.001), moderate vs. large (*p* < 0.001). There was also a significant main effect of “site” (*F*_(2, 4016)_ = 139.95, *p* < 0.001, *η_p_*² = 0.07), demonstrating that LoSH ratings differed across three body regions. Specifically, pain located in the upper (*p* < 0.001) and lower limbs (*p* < 0.001) elicited higher LoSH ratings compared to the lower back. The separate LMM addressing the widespread patterns of PD within the lower leg showed main effect of a “pattern” (*F*_(2, 1004)_ = 174.50, *p* < 0.001, *η_p_*² = 0.26). Namely, two selected discrete patterns -W2 and W3- of pain distribution had higher LoSH values than contiguous one W1 (*p* < 0.001). Results of additional model with between-subject factor “pain status” (having pain vs. not having pain) showed no differences, either with or without the confounding factors, i.e. age, education level, device type, and weight **(Appendix S9)**. Models with “size” × “pain status” interactions were not significant, with and without confounders, for either outcome (LoSH and LoTM; **Appendix S9**), indicating that the LoSH gradients were not affected by having pain.

**Figure 4.**
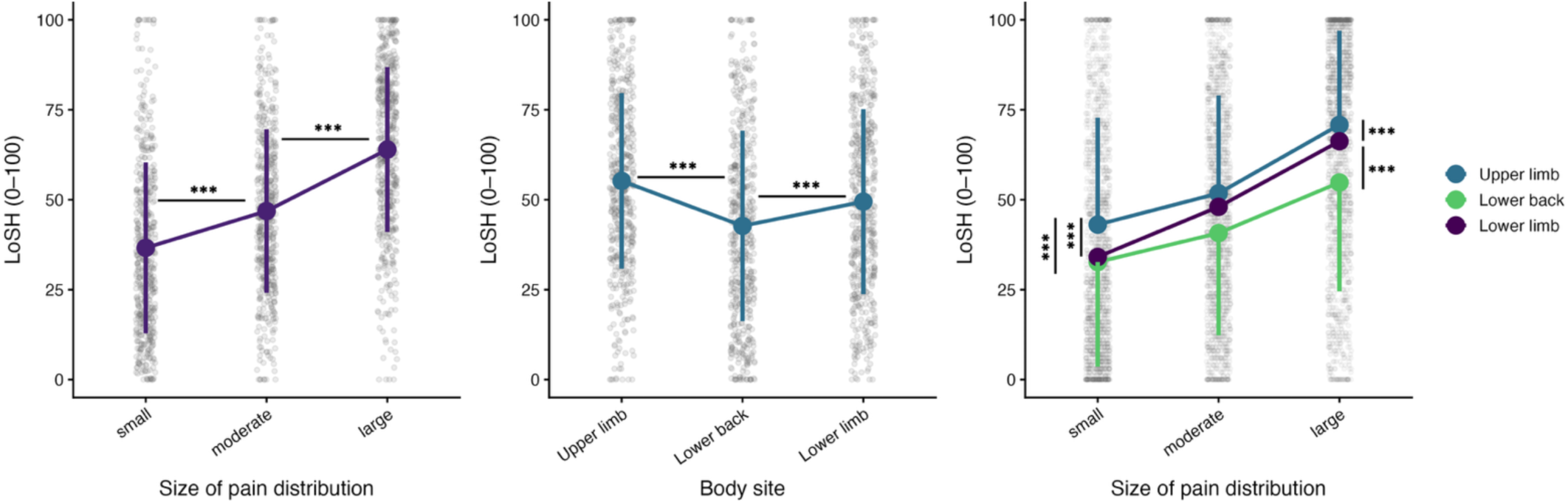
Effects of pain distribution on likelihood of seeking help (LoSH) ratings. Left: Significant gradient in LoSH as a function of pain extent. Middle: A significant but modest effect of body region. Right: Two-way interaction showing higher LoSH ratings for the upper limb when larger pain extents were presented. ***p_(FDR)_ < 0.001.

Similarly to PD, analysis of LoSH of “pain intensity” showed significant “site” × “pain intensity” interaction (*F*_(4, 4016)_ = 4.64, *p* < 0.001, *η_p_*² < 0.01) mainly driven by the ratings of upper limb, which were consistently higher compared to lower limb (*p* < 0.001) and back (*p* < 0.001) for the highest intensity level **(Figure 5)**. Also, significant main effect of “pain intensity” (*F*_(2, 4016)_ = 3950.96, *p* < 0.001, *η_p_*² = 0.66, **Figure 5**) was observed indicating that pain presented with a higher intensity level led to higher reported likelihoods for seeking help: low vs. moderate (*p* < 0.001), moderate vs. high intensity (*p* < 0.001). Significant effects of “site” (*F*_(2, 4016)_ = 13.10, *p* < 0.001, *η_p_*² = 0.01) showed that LoSH ratings were different across three body regions. Specifically, pain located in the lower back elicited lower LoSH ratings compared to the lower (*p* < 0.01) and upper (*p* < 0.001) limb. The separate LMM addressing the effect of pain intensity presented with widespread (discrete) PD patterns also showed significant effect of “pain intensity” (*F*_(2, 1004)_ = 1395.96, *p* < 0.001, *η_p_*² = 0.74). Namely, discrete pattern (W3) of PD (**Figure 1)** had higher LoSH when presented with higher intensity of pain (*p* < 0.001). Analogous analyses conducted on LoTM revealed similar pattern of results and additional tests for potential role of the between-subject factor “pain status” showed no differences **(Appendix S10)**. Models with “intensity” × “pain status” interactions were not significant, with and without confounders, for either outcome (LoSH and LoTM; **Appendix S9**), indicating that the LoSH and LoTM gradients were not affected by having pain.

**Figure 5.**
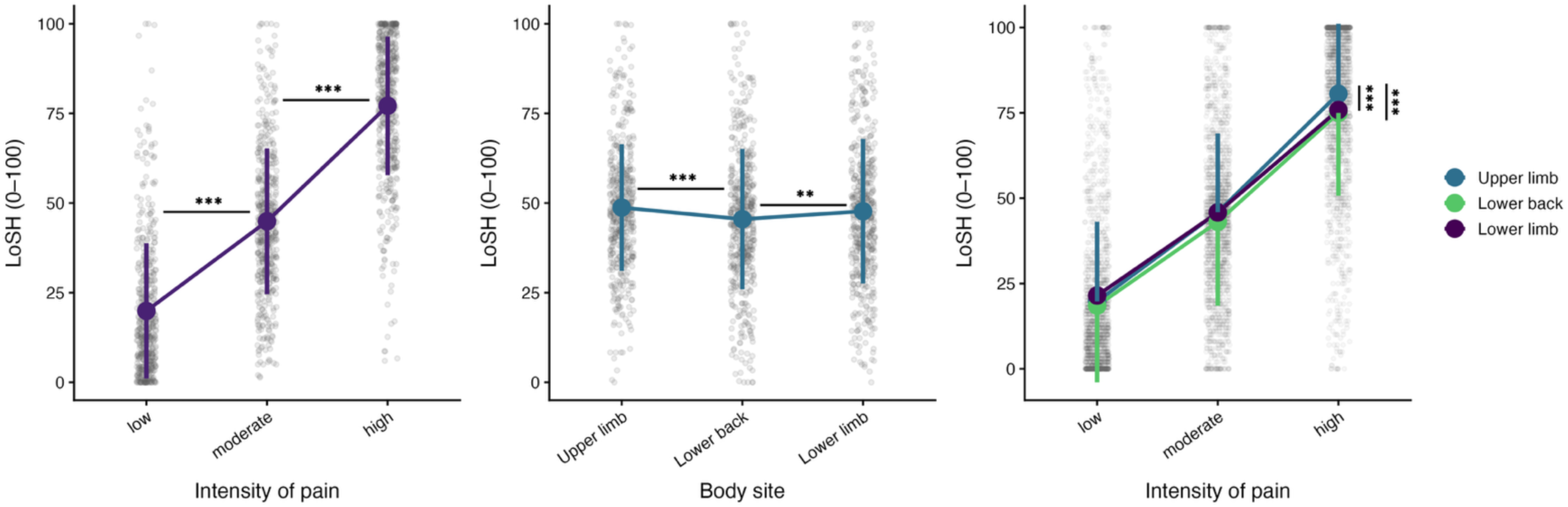
Effects of pain intensity level on likelihood of seeking help (LoSH) ratings. Left: Significant gradient in LoSH as a function of pain intensity: low (2), moderate (5), high (8 out of 10 on Numeric Rating Scale). Middle: A significant but modest effect of body region. Right: Two-way interaction showing higher LoSH ratings for the upper limb when high pain intensity was presented. \*\*\**p*_(FDR)_ < 0.001.

### 3.3. Thought experiments: LoSH versus LoTM

For both outcomes, the “predictor” × “level” interaction was highly significant (LoSH: *F*_(2, 2510)_ = 307.28, *p* < 0.001, *η_p_*² = 0.20, LoTM: *F*_(2, 2510)_ = 304.44, *p* < 0.001, *η_p_*² = 0.20), indicating that the relative influence of pain distribution versus intensity shifted systematically across pain levels. At low pain levels, size/distribution ratings were significantly higher than intensity ratings for both LoSH (*p* < 0.001) and LoTM (*p* < 0.001), suggesting that when pain is mild, its spatial extent is a stronger driver of health-care seeking decisions than its intensity **(Figure 6, Appendix S11)**. At the interim level, the two predictors did not differ for LoTM (*p* = 0.996) and showed difference for LoSH (*p* = 0.025), indicating a crossover point where the two predictors carry approximately equal weight. At high pain levels, the pattern reversed: intensity ratings significantly exceeded distribution ratings for both LoSH (*p* < 0.001) and LoTM (*p* < 0.001), indicating that at severe pain, intensity becomes the dominant predictor of health-care seeking decisions.

**Figure 6.**
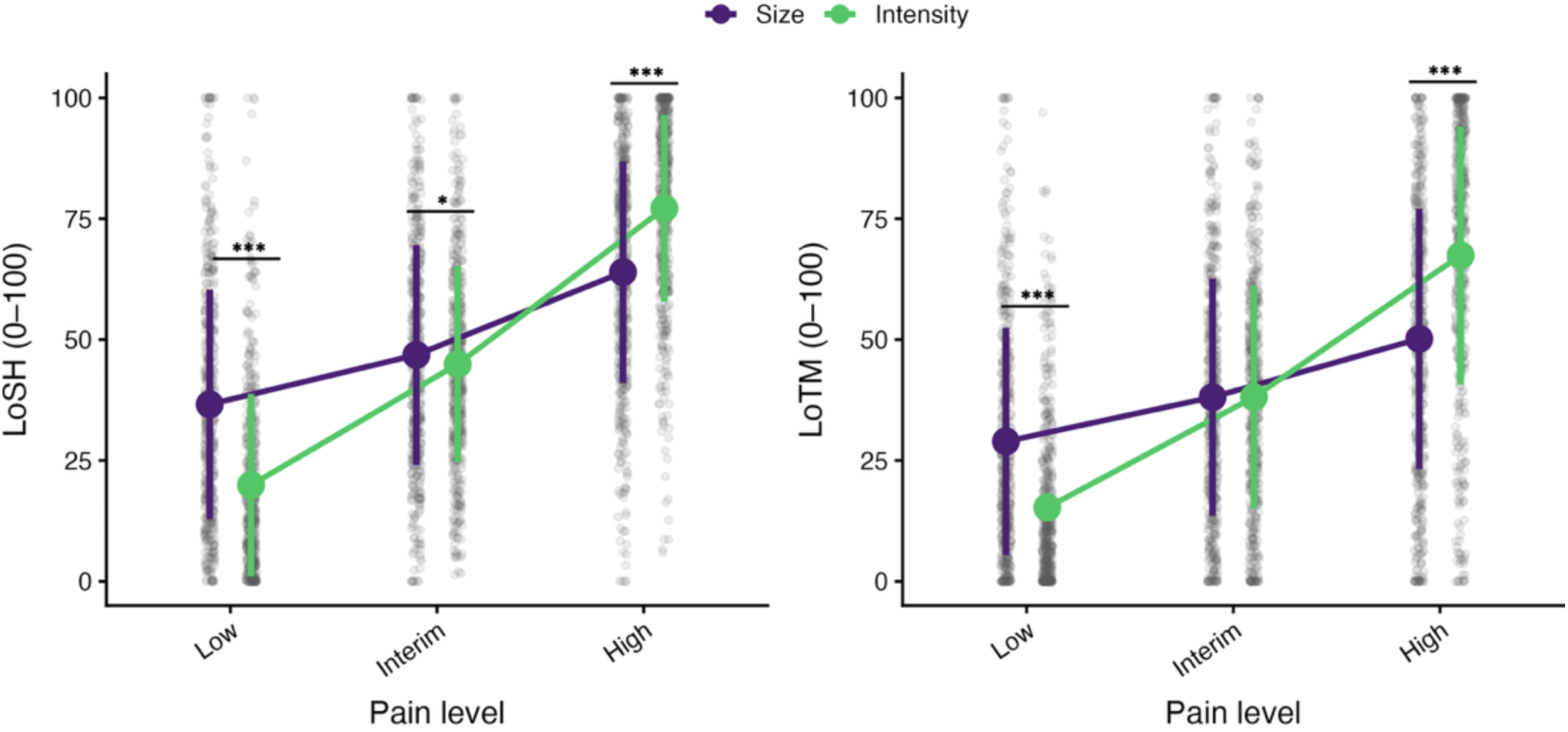
Comparisons of reported likelihoods between pain intensity and pain distribution across all three levels. Plot showcases significant interaction between the “outcome” (intensity vs. pain distribution) and level (mild, interim, high) within likelihood of seeking help (LoSH) and likelihood of taking medication (LoTM). Pain distribution is the stronger predictor of help-seeking and medication-taking at low pain levels, the two predictors converge at interim levels, and pain intensity dominates at high pain levels. \*\*\**p*_(FDR)_ < 0.001.

### 3.4. Trading pain distribution with pain intensity

The GLMM model showed strong trade-off pattern in both tasks **(Figure 7)**. In the “i20 task” (20% intensity reduction fixed while PD reduction varied), increasing the offered PD reduction substantially increased the probability of choosing reduction of that pain dimension (*β* = 0.18 [95% CI: 0.16, 0.19] per 1% extent, SE = 0.01, *z* = 19.94, *p* < 0.001). Expressed as an odds ratio (OR), each additional 10% extent reduction increased the odds of choosing area reduction by a factor of 5.78 (95% CI [4.87, 6.87]). The model-based indifference point (P = 0.50) occurred at 68.2% extent reduction (95% CI [65.4, 70.9]), i.e., participants treated ∼68% reduction in pain extent as roughly equivalent to a 20% reduction in pain intensity. This corresponds to an exchange rate of 1% (intensity) to 3.41% (extent). In the “s80 task” (80% extent reduction fixed while intensity reduction varied), the effect run in the opposite direction **(Figure 7)**. Specifically, increasing the offered intensity reduction decreased the probability of choosing the fixed 80% area reduction option (*β* = −0.10, 95% CI [−0.11, −0.09] per 1% intensity, SE < 0.01, *z* = -21.84. Equivalently, OR per +10% intensity = 0.36, 95% CI [0.33, 0.40]). The model-based indifference point was 28.9% intensity reduction (95% CI [26.0, 31.70]), i.e., participants treated an ∼29% intensity reduction as roughly equivalent to an 80% extent reduction, corresponding to 1% intensity ∼ 2.77% extent.

**Figure 7.**
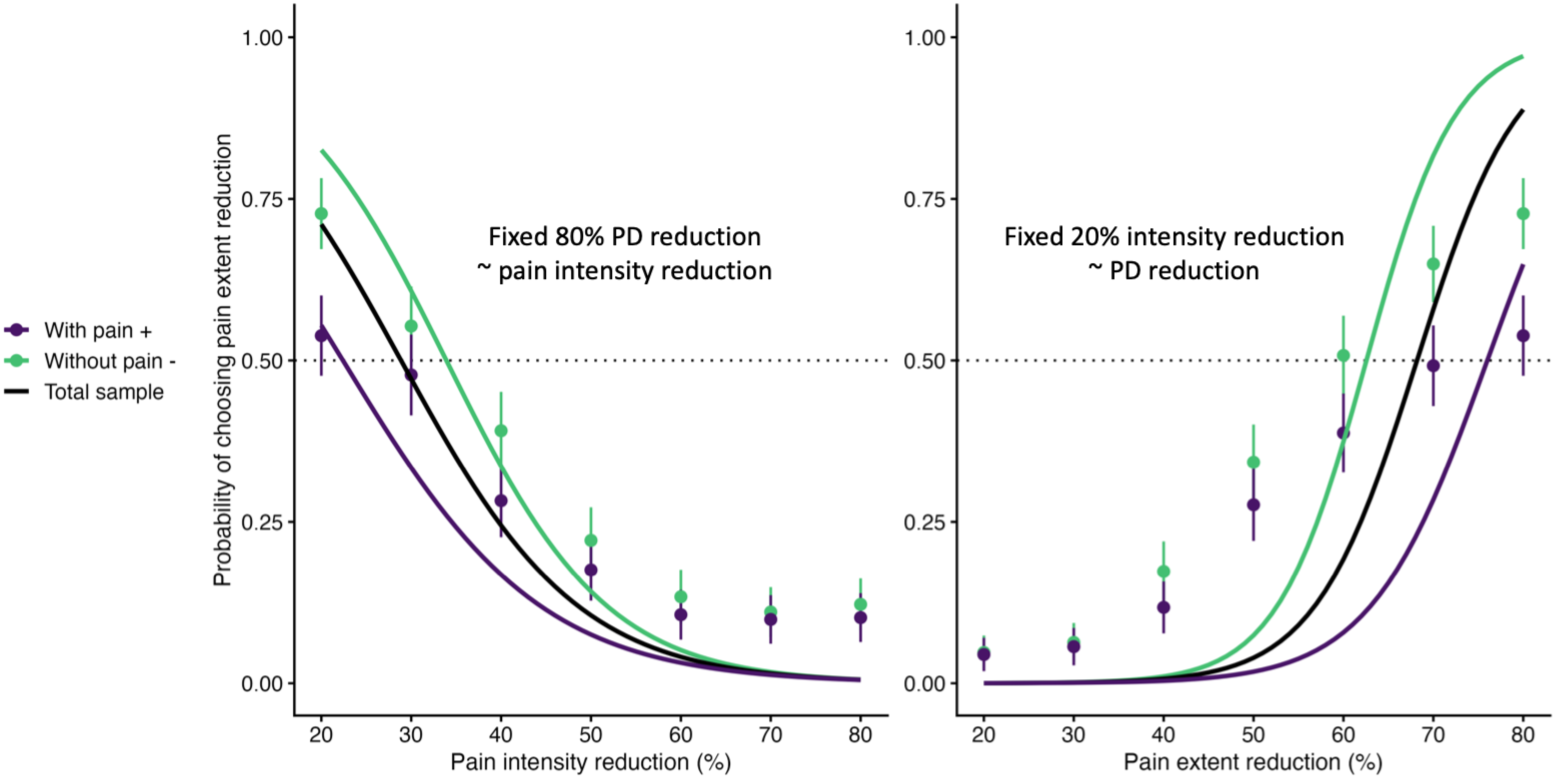
Results of the spatial–intensity trade-off task. Participants traded reductions in pain distribution (PD) against reductions in pain intensity. In one variation of the task, a fixed 80% reduction in PD was compared with varying reductions in pain intensity (left panel). In the other task, a fixed 20% reduction in pain intensity was compared with varying reductions in PD (right panel). Indifference points for the total sample (black) indicated that a 20% reduction in pain intensity was equivalent to a 68% reduction in PD, whereas an 80% reduction in PD corresponded to a 29% reduction in pain intensity. A significant interaction with patient status indicated that participants experiencing pain placed greater value on reductions in pain intensity relative to reductions in PD. Note: group-level probabilities are plotted for visualization with 95% confidence intervals and plotted curves are model- based.

Adding “pain status” as a between-subject factor (having vs. not having pain) revealed a significant “pain status” × “trade-off level” interaction in both tasks (i20: *β* = −0.05, 95% CI [-0.07, -0.02], SE = 0.01, z = -3.54 *p* < 0.001, s80: *β* = 0.02, 95% CI [0.01, 0.04], z = 2.68, *p* < 0.01), indicating that the slope relating trade-off level to choice probability differed between individuals with and without pain. Interpreted qualitatively, the trade-off curve was modestly less steep in individuals with pain on the unified axis. Given this interaction, the main effect of “pain status” (i20: *β* = −1.47, 95% CI [-2.44, -0.51], *z* =-2.99, *p* < 0.01, s80: *β* = −0.71, 95% CI [-1.27, -0.14], *z* = -2.46, *p* < 0.05) should be interpreted as a conditional effect: it reflects the shift in choice probability between pain-status groups at the midpoint of the trade-off scale (trade-off level = 50), rather than a uniform shift across all trade-off levels. All effects remained significant after controlling for the confounding variables, i.e. age, education level, device type, and weight. Group differences were also reflected in the subject-level probability of selecting PD reduction, which differed between individuals with pain and pain-free individuals in both tasks. Namely, the probability of selecting PD reduction over pain-intensity reduction was higher in individuals without pain than in individuals with pain in both tasks (i20: *t*_(497.85)_ = 3.33, *p* < 0.001, s80: *t*_(498.84)_ = 2.66, *p* < 0.01).

## 4. DISCUSSION

In a series of thought experiments, we examined whether variations in PD influence health-related decision-making beliefs, and how these effects compare to those driven by changes in pain intensity. Larger PD and its anatomical location were independently associated with higher LoSH and LoTM. Although the effect of pain intensity was somewhat stronger, the magnitude of PD-related effects was comparable. Importantly, results from two trade-off tasks revealed a systematic relationship between reductions in pain intensity and PD. On average, participants required approximately a threefold larger reduction in PD to consider it equivalent to a given reduction in pain intensity. This “1-to-3” exchange rate was shifted in individuals reporting current pain, who placed relatively greater value on reductions in pain intensity. Together, these findings suggest that spatial characteristics of pain also shape health-related decisions.

Since the emergence of modern pain assessment, the role of PD appeared to be gradually marginalised without a clear rationale. When one of the first clinimetric tools in pain research, the McGill Pain Questionnaire [51], was introduced, a two-dimensional body chart was included as a component of the assessment, but primarily as a supplementary element among many other descriptors of pain - information from the body chart did not contribute to the questionnaire’s score. Today, in the digital era, the possibilities for characterising PD are far more extensive, particularly through digital body-mapping tools [11]. These approaches allow detailed quantification of PD although such methods are not free of limitations. They rely on patient engagement and accuracy in reporting PD (and pain localization) and may require a certain degree of spatial recognition or drawing ability.

Despite these constraints, a substantial body of evidence indicates that PD is not merely a spatial descriptor but also a clinically meaningful index [31,50]. For example, the perceived depth of pain may reflect the involvement of different nociceptive fibers [52], patterns of PD may help distinguish neuropathic from nociceptive pain [28,29] and the overall spatial extent of pain may reflect the presence of central sensitization [47]. Furthermore, to some degree, extent of neural activity within the human neuroaxis can be inferred from the pain extent [23,58]. Given these potential utilities, it is striking that spatial characteristics of pain remain underreported in many clinical studies [67] and are not even part of core outcome set recommendations [20]. Recent commentary has highlighted this gap and argued that pain drawings represent a simple, yet informative tool that could easily be incorporated into routine clinical assessment [12]. Beyond these established clinical uses of PD, our present study addressed a largely unexplored question: How meaningful is PD for the general population when considering pain as a symptom, particularly in comparison with pain intensity? As expected, participants reported gradients of LoSH and LoTM with higher scores assigned to conditions with higher level of pain intensity. The highest ratings of both outcomes occurred for the NRS of 8/10. These findings confirm that pain intensity functions as a primary signal for action in health-related decision-making.

Crucially, PD produced a highly similar decision gradient. When pain intensity was held constant but PD progressively increased, participants also reported a greater LoSH and LoTM. In this sense, PD appears to function as an additional heuristic, guiding responses (decisions) to pain. This observation suggests that PD may carry important societal meaning when individuals evaluate the seriousness of pain. These findings are also consistent with mechanistic insights from experimental pain research. The number and spatial arrangement of nociceptive inputs influence perceived pain intensity through spatial summation of pain (SSP) [2,4,9,14,27,32,59–61] and through interactions with innocuous inputs, as in the thermal grill illusion [15,22,25,53]. Spatial summation has been demonstrated across multiple stimulus modalities and experimental paradigms in which increasing the spatial extent of stimulation under constant intensity leads to reported pain of higher intensity [2,42,59]. In addition, SSP can occur even when stimuli are discrete and spatially separated [4,59,64]. Experimental SSP has also been reported to differ in clinical pain populations: fibromyalgia [37], lateral epicondylalgia [36], osteoarthritis [32] and neuropathic pain [1] showed facilitated summation. Interestingly, several studies, including the present dataset, suggest that the spatial extent of clinical pain correlates with reported pain intensity [5,30,40,48,70,73], echoing some form of SSP. It has also been shown that individuals expect more intense pain from nociceptive stimulation applied to a larger area of the body [56]. These associations raise the possibility that processes related to SSP may operate not only at the peripheral level of nociceptive input [59] or spinal cord [9] but also at the higher level of the neuroaxis where pain perception emerges [56,57]. If so, PD and pain intensity may be tightly interrelated dimensions of pain perception suggesting that pain intensity and PD are not independent features of the pain experience. Instead, they may interact and jointly shape both pain perception and health-care-related decisions. From a clinical perspective, this suggests that interventions targeting not only pain intensity but also the spatial characteristics of pain could potentially enhance treatment effectiveness. This interpretation is consistent with our survey results, in which 61% of participants reported that spatial aspects of pain should be addressed in treatment alongside pain intensity.

Complementing these findings, the results of the SITT tasks constitute a novel attempt to apply a discounting-like methodology [68] to survey research on pain perception. In these tasks, participants were required to trade reductions in pain intensity against reductions in PD, or vice versa. By fitting a binomial model to the choice responses, the indifference point corresponding to a probability of 0.5 was estimated, at which the likelihood of choosing either option was equal. This approach allowed to quantify the relative value assigned to reductions in the two domains at group level. When data from both trade-off tasks were combined, the results indicated a threefold larger reduction in PD to reach equivalence within the intensity domain. Interestingly, participants who reported experiencing pain during the survey, showed a shift in this relationship, suggesting that the perceived cost of reducing pain intensity was higher in individuals with ongoing pain compared with those without pain. Although causal inference is not possible due to the cross-sectional design, these findings suggest that the presence of pain modulates the trade-off between PD and pain intensity, effectively altering their relative exchange. Future work could thus explore this interaction in target populations with widespread vs. local pain. It is important to note, however, that the SITT paradigm probed relative preferences between two domains of symptom reduction, which represents a construct distinct from actual behaviour of choosing treatment reduction in clinical context. Future research should also extend these findings by examining how clinicians perceive and integrate spatial characteristics of pain into their decision-making processes. While the present study focused on lay beliefs and preferences, understanding whether and how healthcare providers weigh pain distribution relative to intensity in clinical assessments and treatment planning could yield valuable insights. This could be accompanied by addressing some limitations of the current work, such as fixed order of items display, which could introduce anchoring effects, whereby earlier items could have shaped participants’ interpretation and evaluation of subsequent items. Future studies should also include measures of healthcare access and healthcare attitudes to better isolate their role in pain-related decision-making as these variables were not measured in the current experiment.

From a broader perspective, our findings carry several implications for both research and clinical practice. First, they suggest that spatial characteristics of pain should be systematically incorporated into pain assessment frameworks. Despite the widespread use of body charts in clinical settings, quantitative information about PD is still rarely reported in clinical trials or epidemiological studies. The increasing availability of digital pain drawings and automated analysis tools now makes it feasible to quantify PD with high precision and minimal burden to patients or clinicians. Second, greater attention to PD may improve phenotyping of pain conditions. Patterns and extent of pain distribution have already been shown to provide diagnostic and prognostic information in several disorders and incorporating PD metrics into predictive models may enhance the ability to identify clinically meaningful subgroups of patients. Third, from a mechanistic perspective, PD may represent an accessible marker of spatial integration processes within the distributed nociceptive system [13]. Integrating experimental paradigms of spatial summation with clinical assessments of PD could therefore provide new insights into how spatial aspects of nociceptive processing contribute to pain perception and chronic pain mechanisms.

In conclusion, individuals do attend to the spatial aspects of pain. The pattern and extent of PD systematically influenced decision-making, such that both seeking medical help and taking analgesics were more likely when pain affected larger body areas. Participants, regardless of pain status, consistently used PD as a relevant cue when evaluating the need for action, and many further endorsed the importance of considering pain distribution alongside pain intensity in pain management. These findings suggest that spatial characteristics of pain contribute meaningfully to how individuals interpret the severity of bodily symptoms. Increasing the visibility of PD as a clinically and mechanistically relevant dimension of pain may therefore improve both assessment and treatment strategies.

## Data Availability

Data supporting the present study are publically available: https://osf.io/c9b3f/

## 5. ACKNOWLEDGEMENTS

The authors have no conflicts of interest to declare. Language editing during writing process was supported by the AI tool (ChatGPT) and components of the Figure 1 were generated by AI. Code was validated and checked for consistency using Claude. The data and code are deposited on OSF.io platform: https://osf.io/c9b3f/. Funding: DFG grant no. 574729638.

