## Appendix file for "Beyond intensity: Pain distribution shapes healthcare- and treatment-seeking beliefs in individuals with and without clinical pain"

*Appendix (Supplementary Materials)*

### S1. Distribution of responses over the data collection period and heat maps

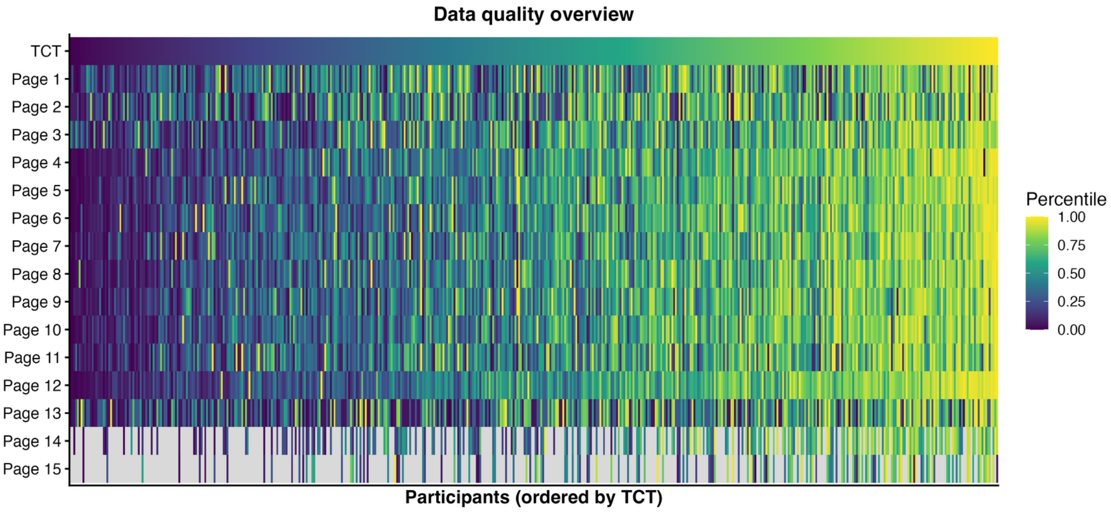


***Heat maps showing individual total completions times (TCT) and times to complete individual pages.*** *Right side of the spectrum indicates faster responders.*

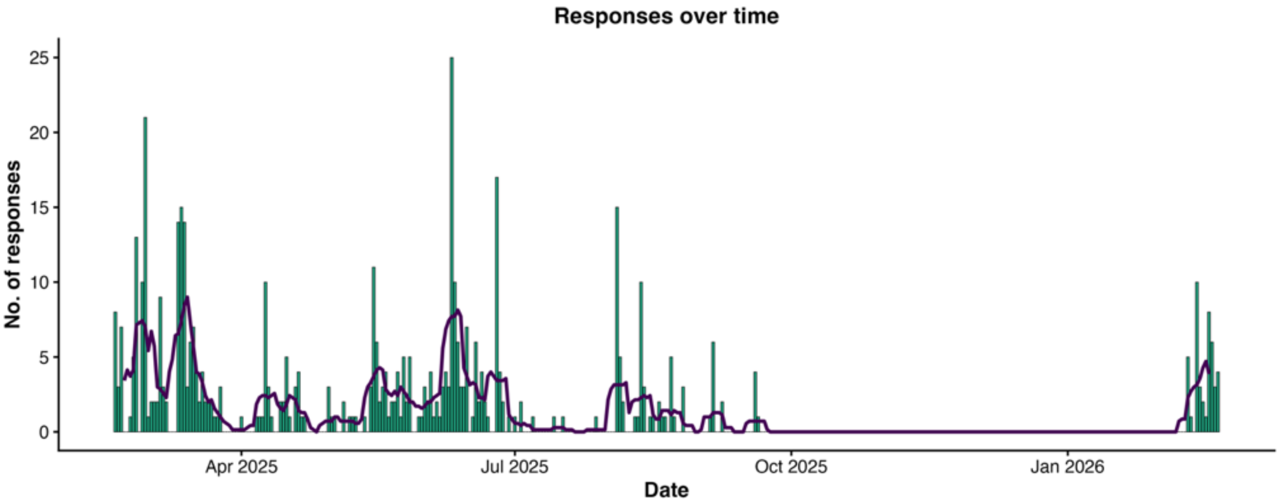


***Total number of completed datasets collected over the study period****. The green bars represent the number of responses collected on each day between February 2025 and February 2026. The purple line depicts the smoothed trend of daily responses. A total of 503 completed datasets were collected over this 12-month period.*

### S2. Piloting phase and study development

Following initial survey development, a two-stage piloting process was conducted to refine the study design and procedures. In the first round, the survey was tested internally within the research team (n = 11). In the second round, it was administered to an external pilot sample (n = 8), comprising individuals with pain (n = 2), individuals without pain (n = 2), and experts in the field (n = 4). Based on structured feedback from both rounds, several modifications were implemented. These included the addition of automated reminders on each survey page to minimize missing responses and the introduction of Likert-scale response options for previously dichotomous attitudinal items assessing beliefs about the relationship between pain characteristics (e.g., spatial extent, depth) and injury severity.

A question assessing the approximate onset date of the current pain episode (MM.YYYY format) was added, and the comparative rating scale evaluating the perceived relative threat of pain extent versus pain intensity was expanded to a symmetrical 11-point format (5–0–5). 11-point Numeric Rating Scales (0–10) embedded in image-based items were standardized to ensure homogeneity across questions. A smartphone-optimized version of the survey was developed to improve usability across devices, and a question assessing educational level was also included to enhance sample characterization. In addition, several semantic refinements were made to improve clarity and precision of items wording, and one subsection of the image-based items (neck area) was removed to reduce time commitment to complete the study.

### S3. Transparency statement

The preregistration stated that exploratory correlation, regression, and predictive modelling analyses would be conducted, including logistic regression models to predict group classification (pain-free vs. pain) and multivariate regression models examining preferences for reducing pain extent versus pain intensity. These analyses were ultimately not performed and are not part of this publication. Following completion of the preregistered primary analyses, we determined that the planned exploratory models were not essential for addressing the primary study aims and would substantially increase the number of statistical tests performed. Instead, we focused on the preregistered primary analyses and a limited set of additional analyses that were more closely aligned with the study objectives and the repeated-measures structure of the data. These additional analyses emerged during peer-review process and included generalized linear mixed-effects models (GLMMs) with covariates, assessments of potential clustering effects, direct comparisons between key outcome variables, and post-hoc power analyses, and additional linear mixed models testing interactions between “size / intensity” and “pain status” variables. This deviation from the preregistered analytical plan is reported here to ensure transparency.

In addition, a novel SITT task was employed, the implementation and analysis of which were not described in sufficient detail in the preregistration protocol. The SITT data were analyzed using GLMMs with a binary response variable indicating whether a pain distribution was selected (0 = no, 1 = yes). Models were fitted using a binomial distribution with the following structure: “response” ~ “trade level” (20:20% to 20:80%) + (1 | “subject”).

The survey remained open for approximately one year, rather than the preregistered eight-week period, due to slower-than-anticipated recruitment and challenges in achieving the a priori target sample size. The extended recruitment period was implemented to maximize participation within the available study timeframe.

### S4. Content of thought experiments

The main part of the survey included blocks of questions (thought experiments). Each block addressed a different body area: upper limb, lower limb (shown), lower back, and widespread patterns. Following presentation of the hypothetical case scenario, an image was displayed showing either different pain distribution (PD) patterns (left) or a constant PD with varying pain intensity (right). Apart from widespread pattern, each block included three LoSH and three LoTM questions. Each question was answered on a Visual Analogue Scale (VAS), with the left anchor set to “very unlikely” (0) and the right to “very likely” (100). The scenario and overall instruction given to participants were:“Imagine you have had pain in your foot / lower leg / entire leg for three to four weeks. The image below shows exactly where the pain is located. Click on the horizontal line between the two response options to indicate your rating.” Table S5 below includes the content and design of the main task with distinction into PD and pain intensity domains.

### S5. Thought experiments

**Pain distribution domain**

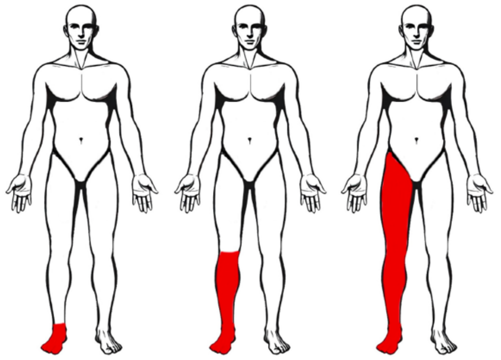

**Likelihood of seeking medical help (LoSH):**

- If you had pain in the red-marked area of the foot (left figure), how likely would you be to take an action and seek medical/therapeutic professional care?
- If you had pain in the red-marked area of the lower leg (middle figure), how likely would you be to take an action and seek medical/therapeutic professional care?
- If you had pain in the red-marked area of the whole leg (right figure), how likely would you be to take an action and seek medical/therapeutic professional care?

**Likelihood of taking pain medication (LoTM):**

- If you had pain in the red-marked area of the foot (left figure), how likely would you be to take a pain medication?
- If you had pain in the red-marked area of the lower leg (middle figure), how likely would you be to take a pain medication?
- If you had pain in the red-marked area of the whole leg (right figure), how likely would you be to take a pain medication?

**Pain intensity domain**

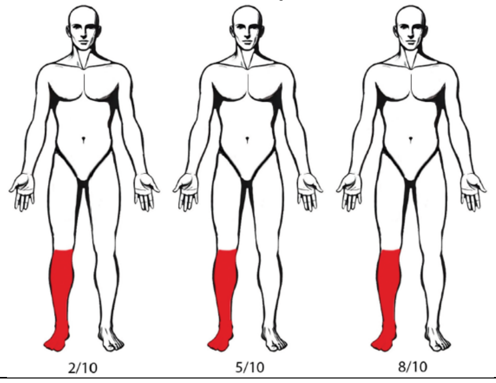


**Likelihood of seeking medical help (LoSH):**

- If you experienced lower leg pain with an intensity of 2/10, how likely would you be to take an action and seek medical/therapeutic professional care?
- If you experienced lower leg pain with an intensity of 5/10, how likely would you be to take an action and seek medical/therapeutic professional care?
- If you experienced lower leg pain with an intensity of 8/10, how likely would you be to take an action and seek medical/therapeutic professional care?

**Likelihood of taking pain medication (LoTM):**

- If you experienced lower leg pain with an intensity of 2/10, how likely would you be to take a pain medication?
- If you experienced lower leg pain with an intensity of 5/10, how likely would you be to take a pain medication?
- If you experienced lower leg pain with an intensity of 8/10, how likely would you be to take a pain medication?

### S6. Group comparisons of total completion time and relative speed index

Total completion time (TCT) in minutes and the relative speed index (RSI) were compared between participants with and without pain using two-sample t tests. Participants in the pain group (n = 248) took significantly longer to complete the survey than participants without pain (n = 255): M = 15.06 min, SD = 6.35 versus M = 11.2 min, SD = 4.71, $t_{(455.49)} = -7.66$, $p < 0.001$. The effect size was moderate to large (d = 0.686), consistent with the additional pain-related items in the survey administered to this group. For the RSI, participants in the pain group had a slightly lower mean RSI than participants without pain: M = 1.08, SD = 0.43 versus M = 1.17, SD = 0.43, $t_{(500.87)} = 2.28$, $p = 0.02$. The effect size was small (d = -0.20), indicating that although the difference reached statistical significance, the magnitude of the group difference in RSI was modest. Together, these results indicate that the longer absolute completion time observed in the pain group is consistent with the additional survey content specific to this group, whereas the small difference in RSI suggests that participants with pain did not respond at a markedly different relative pace once normalized against the cohort median.

**S7. Assessment of clustering of responses at 0 and 100 anchors**

| Outcome | Predictor | N | 0 values | 0 values (%) | 100 values | 100 values (%) |
|---|---|---|---|---|---|---|
| LoSH | size | 4527 | 192 | 4.24% | 291 | 6.43% |
| LoTM | size | 4527 | 385 | 8.50% | 188 | 4.15% |
| LoSH | Intensity | 4527 | 265 | 5.85% | 339 | 7.49% |
| LoTM | intensity | 4527 | 521 | 11.51% | 252 | 5.57% |

**S8. Post-hoc power analyses per reviewer request**

| Outcome | Predictor | Power | 95% CI | n | α |
|---|---|---|---|---|---|
| LoSH | size | 100% | 99.52-100% | 1000 | 0.05 |
| LoTM | size | 100% | 99.52-100% | 1000 | 0.05 |
| LoSH | Intensity | 100% | 99.52-100% | 1000 | 0.05 |
| LoTM | intensity | 100% | 99.52-100% | 1000 | 0.05 |

**S9. Controlling for confounds addressing Hypothesis iii**

To examine whether having pain influenced the outcomes independently of the experimental manipulation, we conducted additional analyses. First, we added "pain status" (having pain vs. not having pain) as a between-subjects factor to the primary LMM ("DV" ~ "pain status" + "body area" × "size" + {1 | "subject"}). Patient status did not significantly predict either LoSH ($F_{(1, 501)} = 0.16$, $p = 0.69$, $\eta^2_p < 0.001$) or LoTM ($F_{(1, 501)} = 0.01$, $p = 0.94$, $\eta^2_p < 0.001$), suggesting that the two groups did not systematically differ in their ratings. Similarly, models in intensity domains showed similar results: LoSH ($F_{(1, 501)} = 0.39$, $p = 0.53$, $\eta^2_p < 0.001$), LoTM ($F_{(1, 501)} = 0.42$, $p = 0.52$, $\eta^2_p < 0.001$).

Second, given that prior analyses revealed group differences in variables: age, weight, screen size of the device, and education, we reran all models including these variables as covariates. Nineteen participants were excluded due to missing covariate data, leaving 484 participants. After controlling for these demographic and clinical variables, the patient effect remained non-significant for all outcomes, and critical factor "size" remained significant for LoSH ($F_{(2, 3864)} = 678.00$, $p < 0.001$, $\eta^2_p = 0.26$), and LoTM ($F_{(2, 3864)} = 437.45$, $p < 0.001$, $\eta^2_p = 0.18$). Similarly, factor "intensity" in intensity models remained significant: LoSH ($F_{(2, 3864)} = 4005.70$, $p < 0.001$, $\eta^2_p = 0.67$), and LoTM ($F_{(2, 3864)} = 3101.67$, $p < 0.001$, $\eta^2_p = 0.62$), consistent with the primary analysis.

Third, we analyzed models with potential interaction terms "pain status" × "size" (e.g., "DV" ~ "pain status" × "size" + "body area" {1 | "subject}), and we did not found any significant interactions: In the size domain, the "pain status" × "size" interaction was not significant for LoSH, either without covariates ($F_{(2, 4018)} = 0.37$, $p = 0.69$, $\eta^2_p < 0.001$) or with covariates ($F_{(2, 3866)} = 0.55$, $p = 0.58$, $\eta^2_p < 0.001$), nor for LoTM, without covariates ($F_{(2, 4018)} = 0.04$, $p = 0.96$, $\eta^2_p < 0.001$) or with covariates ($F_{(2, 3866)} = 0.04$, $p = 0.96$, $\eta^2_p < 0.001$). In the intensity domain, the interaction was likewise not significant for LoSH, without covariates ($F_{(2, 4018)} = 0.58$, $p = 0.56$, $\eta^2_p < 0.001$) or with covariates ($F_{(2, 3866)} = 0.38$, $p = 0.69$, $\eta^2_p < 0.001$), nor for LoTM without covariates ($F_{(2, 4018)} = 2.39$, $p = 0.09$, $\eta^2_p < 0.01$) or with covariates ($F_{(2, 3866)} = 2.41$, $p = 0.09$, $\eta^2_p <$

0.01). None of the eight “pain status” × “size”/“intensity” interactions reached significance (all $p \geq 0.09$), indicating the “size” and “intensity” gradient does not differ between patient and control groups, with or without covariate adjustment.

### S10. Supplementary analyses

Supplementary analyses examined the effects of pain distribution and pain intensity (see below) on the likelihood of taking medication (LoTM) ratings. These analyses revealed response patterns consistent with those observed for the likelihood of seeking help (LoSH) ratings, as reported in the main results section. Consequently, these findings are presented here for reference.

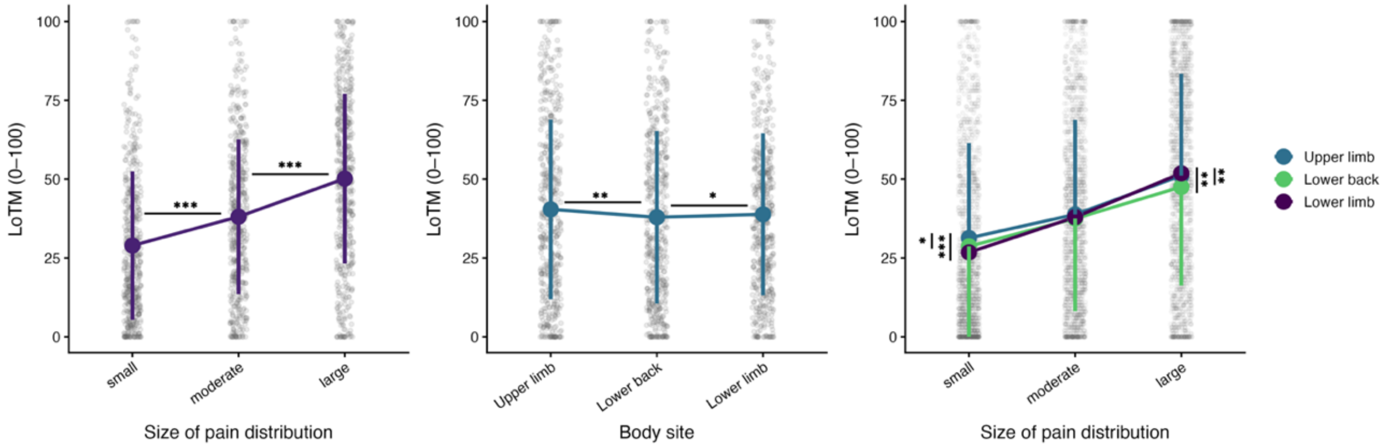


***Effects of pain distribution on likelihood of taking medication (LoTM) ratings.*** *Left: Significant gradient in LoTM as a function of pain extent. Middle: A significant but modest effect of body region. Right: Two-way interaction showing similar patterns in LoTM ratings for all body regions when larger pain extents were presented.* $^{*}p_{(FDR)} < 0.05$, $^{**}p_{(FDR)} < 0.01$, $^{***}p_{(FDR)} < 0.001$.

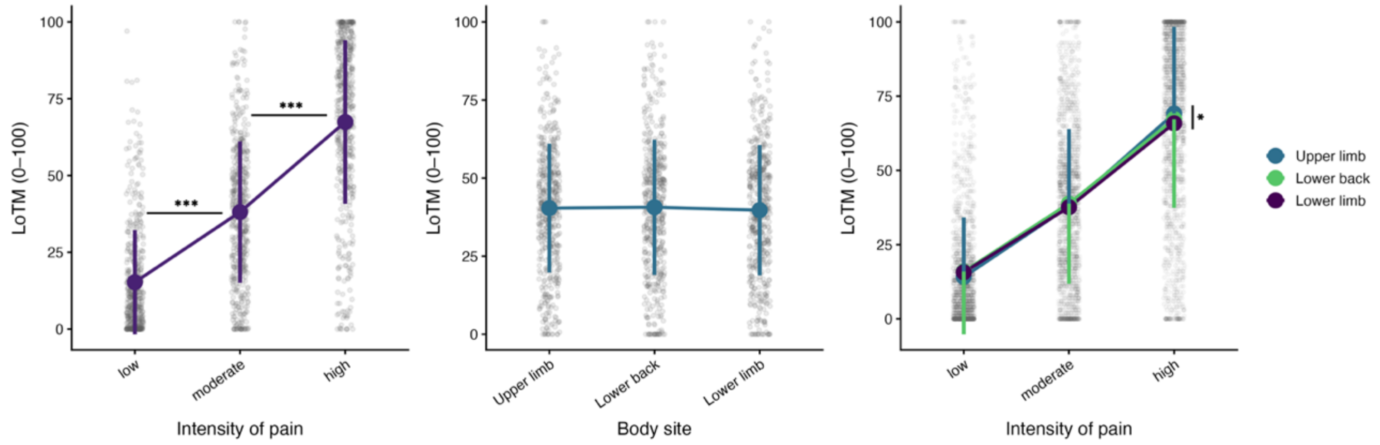


***Effects of pain intensity on likelihood of taking medication (LoTM) ratings.*** *Left: Significant gradient in LoTM as a function of pain intensity. Middle: A non-significant effect of body region. Right: Two-way interaction showing similar patterns in LoTM ratings for all body regions when larger pain intensities were presented.* $^{*}p_{(FDR)} < 0.05$, $^{***}p_{(FDR)} < 0.001$.

S11. Pairwise contrasts: Size vs Intensity at each pain level

| Variable | Level | Mean diff. | SE | df | t | $p_{FDR}$ |
|---|---|---|---|---|---|---|
| LoSH | Low | +16.69 | 0.85 | 2510 | 19.59 | $< 0.001$ |
| | Interim | +1.92 | 0.85 | 2510 | 2.25 | 0.025 |
| | High | −13.19 | 0.85 | 2510 | −15.47 | $< 0.001$ |
| LoTM | Low | +13.67 | 0.89 | 2510 | 15.40 | $< 0.001$ |
| | Interim | 0.00 | 0.89 | 2510 | 0.00 | 0.996 |
| | High | −17.25 | 0.89 | 2510 | −19.42 | $< 0.001$ |

*Note. Mean difference = Size − Intensity (positive values indicate size rated higher). p-values are FDR-corrected (Benjamini-Hochberg) across pain levels within each dependant variable. Degrees of freedom estimated via Kenward-Roger method. LoSH = Likelihood of Seeking Help; LoTM = Likelihood of Taking Medication.*
